# Patterns of Injury in a Gaza War Hospital

**DOI:** 10.1101/2024.06.27.24309570

**Authors:** Abdalkarim Alsalqawi, Richard Villar

**Author notes:** **Address for correspondence:** Richard Villar, Consultant Orthopaedic Surgeon. **Competing interest statement:** Neither the authors nor their institutions received any payments or services in the past 36 months from a third party that influenced the submitted work. No funding was required, nor provided for this study.

## Abstract

This study describes the patterns of injury observed in a Gaza war hospital, focusing on 110 consecutive patients. Of these, two had conditions unrelated to trauma, so the analysis was of the remaining 108 casualties. No military personnel were seen. The data reveal clear trends, including a high prevalence of explosive injuries (86.36%), a low prevalence of gunshot wounds (8.18%), a notable proportion of female (34.55%) and child (23.64%) casualties, and the occurrence of multiple injuries (1.73 injuries/patient) because of the use of high explosive. There were 187 injuries identified including 128 fractures. Of these fractures, 64.84% were of the lower limb and 28.91% of the upper limb. Of the 128 fractures, 79 (61.72%) were clinically and/or radiologically infected. The most frequently infected fracture was the compound tibial and fibular fracture, which showed an infection rate of 92.86%. These findings highlight the unique and tragic nature of the Gaza conflict, the increasing injuries to civilians, including women and children, and the long-term healthcare that will be needed.

## Introduction

Conflict zones present unique medical challenges, with injury patterns often reflecting the nature and intensity of the violence. The precise recording of data is difficult, sometimes impossible, when warfare is ongoing.

Immediately after a 7 October 2023 Hamas-led attack on Israel, Israel began the bombing of Gaza. On 13 October 2024, Israel commenced ground operations in Gaza in the form of small incursions and overnight raids, and on the night of 27 October 2023 it launched a full-scale invasion of the territory. This was called Operation Swords of Iron.

At the time this paper was written (from 21 May 2024), and although precise figures are likely to be inaccurate, it was stated that more than 37,000 people had been killed – 35456 Palestinians and 1478 Israeli. Official data also suggested that 79476 had been wounded and more than 10,000 were still missing under rubble. There had been 450 attacks on healthcare facilities, which had resulted in 723 people killed and 924 injured. There were also 128 health workers in detention. Of Gaza’s 36 hospitals, 32 had been damaged, none was fully operational but 15 (42%) were partially so^i^.

One of these partially operational facilities was the Shuhada al-Aqsa Hospital in the city of Deir-al-Balah, which is one of 15 public hospitals in Gaza and was founded in 2001. It normally contained 166 beds and included a specialist maternity unit. At the time this paper was written, the hospital housed approximately 700 patients, while 3000 displaced Palestinians were in tents in the hospital grounds. The maternity unit had closed, turning itself over to the care of trauma, while an earlier visit by the World Health Organization had found that 70% of staff and patients had fled^ii^. Many staff were working unpaid and on a voluntary basis. There was no regular system of patient allocation to either a ward or consultant. Patients were admitted to whatever portion of the hospital had capacity, and care was frequently provided by family and friends. There was no catering, limited ward cleaning, no prosthetist, scarce rehabilitation, little laboratory support, and minimal ward care other than what could be offered by frequently unqualified nursing skeleton staff. The hospital’s original two operating theatres had been expanded to five by the conversion of three delivery suites into temporary operating theatres. There was regular fighting within two kilometres of the hospital, occasional attacks on the hospital itself, and surveillance drones flying permanently overhead.

This study reports on 110 consecutive civilian patients who occupied one area of the Shuhada al-Aqsa Hospital, an area that would normally contain 40 patients, 20 male and 20 female. The hospital did not treat military patients. The purpose of this survey is to gather a better understanding of the type of patient admitted to what had become a war hospital in a zone of conflict. We regard these data as being essential knowledge for all who may wish to work in such an environment and of value to organisations, charitable or otherwise, who seek to support healthcare provision in the Occupied Territories.

## Methods

We collected data from 110 consecutive in-patients who occupied one area of the Shuhada al-Aqsa Hospital and who had been admitted through the hospital’s Emergency Department. We recorded information on age, gender, type of injury, the number of injuries each patient had sustained, and whether the injury had been compound on admission. We categorised the patients by age groups into (i) under 18 years, (ii) 18-35 years, and (iii) over 35 years. Injury mechanisms were classified as explosive, gunshot wounds, or other injuries.

### Permission

Permission to undertake this study, and report these findings, was given by the administration and Medical Director of Shuhada al-Aqsa Hospital, Deir-al-Balah, Gaza. We have ensured that no patient is identifiable in this study. Because of ongoing conflict, there was no Ethics Board in existence, but we considered this to be unnecessary in any event.

## Results

Of the 110 consecutive patients, there were 26 (23.64%) aged under 18 years, 56 (50.91%) aged 18-35 years, and 28 (25.45%) aged over 35 years. There were 38 (34.55%) females and 72 (65.45%) males.

There were 95 (86.36%) casualties created by an explosion, 9 (8.18%) by gunshot, and 4 (3.64%) had been injured by other means (fall - 3 casualties, torture - 1 casualty). The remaining 2 patients (1.82%) had cellulitis (1 patient) and diabetes (1 patient). All those who had been injured by gunshot were male. See Table 1 for a breakdown of the study group by age, gender, and mechanism of injury.

**Table 1.** The study group (n=110)

|  | **Number**<br>**of**<br>**patients** | **Percentage of**<br>**total (n=110)** |
| --- | --- | --- |
| Age<18 years | 26 | 23.64 |
| Age 18-35 years | 56 | 50.91 |
| Age >35 years | 28 | 25.45 |
| Female | 38 | 34.55 |
| Male | 72 | 65.45 |
| Explosive injuries | 95 | 86.36 |
| Gunshot wounds | 9 | 8.18 |
| Fall | 3 | 2.73 |
| Torture | 1 | 0.91 |
| Other conditions | 2 | 1.82 |

Of the 110 patients, 108 (98.18%) were thus casualties created by the ongoing conflict. These 108 casualties had 187 injuries between them, giving a mean of 1.73 injuries per patient (range 1 to 5). Of the 108 casualties, 59 (54.63%) had more than one injury.

Of the 108 casualties, 87 (80.56%) had sustained injuries to their upper and/or lower limbs with or without fracture, and 21 (19.44%) had sustained injuries confined to other areas. There were 11 (10.19%) casualties with burns. In the 108 casualties, there were 21 (19.44%) amputations.

Eighty-six (79.63%) casualties had sustained injuries to their musculoskeletal system, leading to 128 fractures. The breakdown of the number of fractures as a percentage of these 128 fractures was: tibia/fibula - 42 (32.81%), forearm/elbow/hand - 28 (21.88%), femur - 23 (17.97%), ankle/foot - 11 (8.59%), humerus/scapula - 9 (7.03%), pelvis/hip – 7 (5.47%), skull - 6 (4.69%), zygoma - 1 (0.78%), spine - 1 (0.78%).

Of the 128 fractures, 79 (61.72%) were clinically and/or radiologically infected. Of these infected fractures, 57 (72.15%) were of the lower limb, and 21 (26.58%) were of the upper limb. There was one (1.27%) infected zygomatic fracture. The most frequently infected fracture was the compound tibial and fibular fracture, which showed an infection rate of 92.86%. See Table 2.

**Table 2.** Distribution of fractures (n = 128) in the108 casualties with their total of 187 injuries.

| **Bone(s) fractured** | **Number of fractures (n = 128)** | **Percentage of all injuries (n = 187)** | **Percentage of all fractures** | **Number of infections** | **Percentage infected** |
| --- | --- | --- | --- | --- | --- |
| *Lower limb* |  |  |  |  |  |
| Tibia/fibula | 42 | 22.46 | 32.81 | 39 | 92.86 |
| Femur | 23 | 12.30 | 17.97 | 9 | 39.13 |
| Ankle/foot | 11 | 5.88 | 8.59 | 6 | 54.55 |
| Pelvis/hip | 7 | 3.74 | 5.47 | 3 | 42.86 |
| *TOTAL* | *83* | *44.38* | *64.84* | *57* | *68.67* |
| *Upper limb* |  |  |  |  |  |
| Forearm/elbow/hand | 28 | 14.97 | 21.88 | 18 | 64.29 |
| Humerus/scapula | 9 | 4.81 | 7.03 | 3 | 33.33 |
| *TOTAL* | *37* | *19.78* | *28.91* | *21* | *56.76* |
| *Other* |  |  |  |  |  |
| Skull | 6 | 3.21 | 4.69 | 0 | 0 |
| Zygoma | 1 | 0.54 | 0.78 | 1 | 100 |
| Spine | 1 | 0.54 | 0.78 | 0 | 0 |
| *TOTAL* | *8* | *4.29* | *6.25* | *1* | *12.50* |

In addition to these fractures, wounds were also seen to the following, percentages being quoted as their proportion of the 187 injuries: abdomen - 8 (4.28%), eye - 5 (2.67%), thorax - 4 (2.14%), head - 2 (1.08%). The head injuries were in addition to the skull fractures also seen.

## Discussion

Our study was undertaken at a single moment in time during a period of intense conflict that was directed not only at military personnel but also civilians, healthcare facilities, humanitarian organisations, and others. We have demonstrated some notable differences to what has been previously reported from other zones of conflict. It must be appreciated that there are significant logistic issues when undertaking any type of research during a time of war, so our data should be seen in that light. It should be regarded as a generalisation.

The study group of 110 patients represented approximately 15.71% (110/700) of the actual occupancy of the hospital but is a consecutive series of casualties on two floors of one hospital building, and for which full data capture was possible. In the absence of conflict, there would normally have been 40 patients in that area, 20 male and 20 female. There was no formal admission system in the hospital, casualties simply finding whatever space they could.

Approximately 65% of our casualties were male and 35% were female (see Table 1). This is a slightly higher injury rate for females than reported from Baghdad in 2018 by Shaak et al who undertook a randomised household cluster survey of 900 households. They found 29% of the injuries were in females^iii^. Our higher rate of injury in females may reflect intentional targeting but may also be a feature of the tightly packed environment of Gaza, and its very urban nature, where it is difficult for targeting to be accurate.

Injuries to children have long been regarded as a serious problem, both in Gaza and elsewhere. We found that 23.64% of our study group was aged under 18 years. This was far higher than we were either expecting or hoping to find. McIntyre, in 2020, looked at 38 published papers on the casualties of the Syrian Civil War and found that children comprised 16.1% of cases^iv^. Again, we are uncertain if our finding reflected intentional targeting, or the difficulty of undertaking warfare in an urban environment. Whatever the reason, we consider our finding of 23.64% to be unacceptably high.

Explosive injuries had occurred in 86.36% of our patients while only 8.18% had been wounded by gunshot, and then only in males. Much work has been undertaken on the likely patterns of injury experienced in war zones. Such patterns have varied depending on the conflict studied. Rustmeyer, Kranz and Bremerich^v^ evaluated ten retrospective studies for the period 1982-2005 that covered war injuries from Vietnam, Lebanon, Slovenia, Croatia, Iraq, Somalia, and Afghanistan and found differences in the causes of injuries. Injuries from fragments were more common during the 1990s than during the earlier Vietnam War (1955-1975), where shooting injuries predominated. Research on military casualties in Kuwait during Operation Desert Storm (1991) showed that 48% suffered fragmentation wounds and only 10% sustained gunshot wounds. Meanwhile the Wound Data and Munitions Effectiveness Team (WDMET) data suggest that, in modern conflict, the causes of injury are bullet (23%), fragment (62%), burn (6%), blast (3%), and other (6%)^vi^. Our finding that 86.36% of our patients had been exposed to explosive injuries was far higher than reported by others.

It is difficult to properly compare military with civilian injuries, especially in a location such as Gaza. None of the patients in our study group was military and although dying is the same in both civilian and military arenas, military combat trauma is normally different to that seen in civilians^vii^. Casualty evacuation, for example, is generally quicker for an injured soldier and the fighter alongside the casualty may also have been trained in combat first aid, thereby improving the odds of limiting haemorrhage, plugging a sucking chest wound, or maintaining an airway. None of this applies to the injured civilian. Most reports of wounds concern the experience of the military. A few have looked at conflict-related injuries sustained by civilians and local combatants. For example, Wild et al^viii^ studied 49 reports from conflicts in 18 countries and found blast to be the mechanism of injury in 50.2% of the casualties studied, again far lower than our own finding of 86.36%.

It is our view that the use of high explosive in civilian-dominated surroundings has led to much of what we report here. The high explosive, however accurately targeted, is also indiscriminate. For example, a 500-pound blast will severely damage, injure, or kill everything or anyone within 20 metres (65 feet) while a 2000-pound blast will increase this radius to 35 metres (115 feet)^ix^. High explosive delivered into an inhabited building can be even more destructive.

We recorded 128 fractures in our 108 casualties. Many (93.75%) of these fractures were of an extremity (64.84% lower limb, 28.91% upper limb – see Table 2). This subdivision of fractures into those of the upper and lower limbs matches with work by Burkle et al, albeit on military casualties in Kuwait during Operation Desert Storm (1991). Their research found that extremity wounds occurred nearly twice as frequently in the lower extremities as in the upper extremities^x^. Our study showed that fractures of the lower limb were 2.24 times more common than those of the upper limb, largely agreeing with Burkle et al.

We were alarmed by the high infection rate of the fractures. Of the 128 fractures, 79 (61.72%) were infected. Certain types of fracture were more commonly infected than others. For example, the infection rate of a compound tibial and fibular fracture was 92.86% (see Table 2). Bacterial contamination of these wounds occurs either at the time of injury or during treatment. Much research has been undertaken into this matter as the infection rate of war wounds is well known to be high. For example, Murray et al^xi^ took bacterial swabs from 61 wounds during conflict in Iraq and cultured bacteria in 49% of them. Research during the Croatian Civil War identified that 61% of patients whose fractures had been treated by external fixation, acquired their infection outside hospital, while the remaining 39% became infected while in hospital^xii^. Many of the injuries we saw were compound. For our 108 casualties in Gaza, 71 (65.74%) had compound injuries on admission, created largely by penetrating shrapnel and debris from missile and bomb explosions. Our findings were higher than that reported by the research^xiii^ of Islinger, Kuklo and McHale, which showed 51% of conflict wounds to be compound.

Meanwhile a study of 211 battlefield casualties from Iraq^xiv^ showed 56 (26.42%) to be infected, although infection was most common in blast injuries. Our fracture infection rate of 61.72% was far higher than this, a finding that bodes badly for the future and has significant implications for the long-term care of those casualties in Gaza who have developed, or will develop, post-injury infection and/or osteomyelitis.

The apparent liberal use of high explosive in Gaza created multiple injuries in many of our casualties, and a total of 187 injuries overall. This finding produced a mean number of injuries per casualty of 1.73 (range 1 to 5). Of the 108 casualties, 59 (54.63%) had more than one injury. We consider this important as it means that great care is needed when assessing a fresh casualty after an explosive injury, as the presence of multiple injuries is likely.

The civilian casualty load troubled us throughout the period of this study. It is evident that there has been a change in warfare from military-led cross-border traditional wars^xv^, to those focused on local communities^xvi^ and civilians^xvii^. In traditional wars, soldiers are the primary target and form the main group of casualties. Advances in the arms industry, changes in strategies, in addition to adjustments in ideologies in recent times, have allowed battlefields to move into civilian territory. This has made the civilian population, irrespective of age and gender, more vulnerable to the effects of warfare^xviii^.

Our study has clearly shown this, with the huge civilian casualty load of a frontline Gaza hospital, an establishment that had become a trauma unit through circumstance rather than design. The increased percentage of females, the wounded children, the large number of fractures affecting both upper and lower limbs, the high infection rate, and the multiple injuries created using high explosive, have all contributed to the figures we report here. We have not even considered the evident need for long-term care and the damage being created to mental health. Those are both topics for further research.

Our recommendation is simple. The war must cease.

## Data Availability

All data produced in the present study are available upon reasonable request to the authors

## Acknowledgements

Our sincerest thanks to all at the Shuhada al-Aqsa Hospital in Deir-al-Balah, Gaza, who helped make this research possible, and who were working under almost unimaginable, high-risk circumstances throughout the period of this study. Many have perished or have been displaced. Thank you also to the Palestinian Ministry of Health, to the International Rescue Committee (IRC), and Medical Aid for Palestinians (MAP).

